# Non-Endoscopic Screening for Barrett’s Esophagus Using a DNA Methylation-Based Assay: Real-World Experience in 11,991 Patients

**DOI:** 10.1101/2025.11.20.25340693

**Authors:** Michael S. Smith, William T. Chen, Paul S. Panzarella, Ivy T. Le, Suman Verma, Victoria T. Lee, Lishan Aklog

## Abstract

**Background:** Barrett’s esophagus (BE), defined by specialized intestinal metaplasia (SIM), is the recognized precursor to esophageal adenocarcinoma (EAC). Despite established screening guidelines for high-risk populations, uptake of endoscopic screening remains low. We report the results of early commercial use of a non-endoscopic BE screening strategy, EsoGuard^®^ (EG), a molecular biomarker assay performed on esophageal cell samples collected via a companion swallowable balloon capsule device (EsoCheck^®^, EC).

**Methods:** We retrospectively analyzed the feasibility and tolerability of EG/EC as a non-endoscopic approach to BE evaluation in patients tested from January 2023 through June 2024. Multivariable logistic regression was used to evaluate risk factors associated with positive EG results.

**Results:** Among 11,991 patients tested, 11,355 (94.7%) had successful cell collection with the EC device, with procedures averaging less than 2 minutes and no serious adverse events reported. Over 95% of surveyed patients would recommend the test to others. Overall, 93.1% (10,566/11,355) of collected samples had binary EG results, among which 17.8% had positive assay results suggesting the presence of BE. Positivity rates increased with age, with age >50 years as the strongest individual predictor of a positive EG result.

**Conclusions:** These data represent the largest real-world experience with EG and EC to date, demonstrating excellent safety and tolerability. The findings support the feasibility of large-scale implementation and suggest that EG/EC use may improve access to guideline-recommended BE screening across diverse healthcare settings.

## 1. Introduction

The incidence of esophageal adenocarcinoma (EAC), the most common esophageal cancer in the U.S., has increased greatly over recent decades, with 5-year survival remaining poor at about 20% [1,2]. Barrett’s esophagus (BE), a condition of specialized intestinal metaplasia (SIM) within the tubular esophagus, is the direct precursor to EAC [3,4]. In contrast to the lethality of EAC, BE is treatable endoscopically. Current U.S. gastroenterology guidelines recommend screening in high-risk patients with multiple well-defined risk factors. These include symptoms of gastroesophageal reflux disease (GERD), male sex, white race, age >50 years, obesity, tobacco smoking, and family history of BE or EAC [5,6].

Unfortunately, in current practice, only approximately 10% of eligible patients undergo endoscopic evaluation for BE, an issue that is likely multifactorial in nature [7,8]. Patient failure to recognize risk factors including GERD, limited knowledge of screening recommendations by primary care physicians, and patient concerns regarding the logistics and risks of upper endoscopy (esophagogastroduodenoscopy, EGD) contribute to the low rates of endoscopic screening and sub-optimal disease detection [9–11]. The incidence of BE in the target population is estimated at 5-15%; however, true rates likely are higher due to historically low screening rates [12]. Significant advances have been made over the last decade in development of non-endoscopic biomarker-based tests that can serve as adjunctive tools for the diagnosis of BE. The 2022 guideline updates from the American College of Gastroenterology (ACG) and clinical practice updates from the American Gastroenterological Association (AGA) describe “minimally invasive non-endoscopic swallowable cell collection device(s) combined with biomarkers” as an alternative to endoscopy for BE screening in high-risk patients [5,6]. Within this paradigm, non-endoscopic cell collection and biomarker-based testing serves as an initial in-office triage step, directing patients to either (a) medical management if test results are negative or (b) EGD with or without biopsies if positive, for confirmation and histopathologic staging.

The first commercially available molecular biomarker test in the U.S. to support the diagnosis of BE is the EsoGuard^®^ (EG) Esophageal DNA Test (Lucid Diagnostics Inc., New York, NY). EG uses targeted next-generation sequencing (NGS) to detect patients with cellular changes associated with BE. The assay is performed on distal esophageal cell samples collected non-endoscopically using EsoCheck^®^ (EC), an FDA 510(k)-cleared swallowable tethered balloon-capsule device (K230339; Lucid Diagnostics Inc., New York, NY), which can be administered in-office without sedation. This assessment strategy is intended to increase BE screening uptake and disease detection in high-risk populations without disproportionate use of endoscopic resources. We report 18 months of real-world feasibility and tolerability data from nearly 12,000 consecutive patients who underwent EG/EC as a first stage of clinically indicated BE screening.

## 2. Materials and Methods

### 2.1. Data Collection and Participants

A retrospective cross-sectional analysis was conducted including patients who underwent EC esophageal cell collection and EG biomarker testing to screen for BE between January 2023 and June 2024. Ethics approval was obtained for the study plan through the Western Institutional Review Board (WIRB®; study number 20244309), with the Board finding the study meets requirements for waiver of consent under 21 CFR 50.22. Consecutive patients who underwent EC administered by Lucid Diagnostics personnel were included, with baseline characteristics and procedure data recorded in the central laboratory database as part of standard practice. Standard operating procedures for EC device administration training were followed, including a minimum of 10 proctored cell collections to demonstrate basic competency before personnel were allowed to perform independent (non-proctored) cell collections. Data for individuals who underwent testing performed by their own physicians or affiliated staff were not readily available in a deidentified manner and therefore were not included in this analysis.

Patient demographics, BE risk factors, details of the EC cell collection procedure (namely procedure duration, patient gag response, successful vs. unsuccessful collection, and any adverse events), and EG results were acquired from the central laboratory database in an aggregate fashion without specific patient identifying information. Although presence or absence of GERD symptoms was captured in the database, duration of symptoms was not. Results from a voluntary patient survey on the cell collection experience also were obtained from the central laboratory database. The patient’s gag response (GR) during cell collection was assessed by the EC device administrator on a five-point scale (1 = minimal to no gag, 5 = unable to complete the cell collection) and detailed in *Appendix Supplemental Table 1*.

As an FDA 510(k)-cleared device, issues pertaining to patient safety, device malfunctions/failures, and usability issues for EC have been tracked in compliance with Medical Device Reporting (MDR) requirements. Event documentation was performed in accordance with Lucid Diagnostics’ manufacturing and quality management systems (QMS) requirements. Data pertaining to EC safety during the study timeframe also was collected from the QMS system.

### 2.2. EsoCheck (EC) Esophageal Cell Sample Collection

The EC device is indicated for use in non-endoscopic collection and retrieval of surface cells of the esophagus in the general population of adults and adolescents, 12 years of age and older [13]. The balloon capsule features Collect + Protect technology™ for targeted sampling (Figure 1). The device is swallowed in encapsulated form, after which the balloon is inflated by passing air through the hollow tether using a syringe. Gentle pressure on the tether generates cell sample collection along the textured surface of the inflated balloon, as it is withdrawn through the distal 5-7 cm of the esophagus. The balloon is then syringe-deflated into the capsule prior to device retrieval, protecting the cell sample from dislodgement and contamination in the upper esophagus and oropharynx. This EC cell collection process follows the device instructions for use and has been described in previous publications [14–16]. Administration of EC can be performed by any trained personnel, including physicians and non-physician healthcare providers such as nurse practitioners, registered nurses, and medical assistants. Collection can occur in any office setting, including a clinic, at health fairs, or at designated commercial test centers.

**Figure 1.**
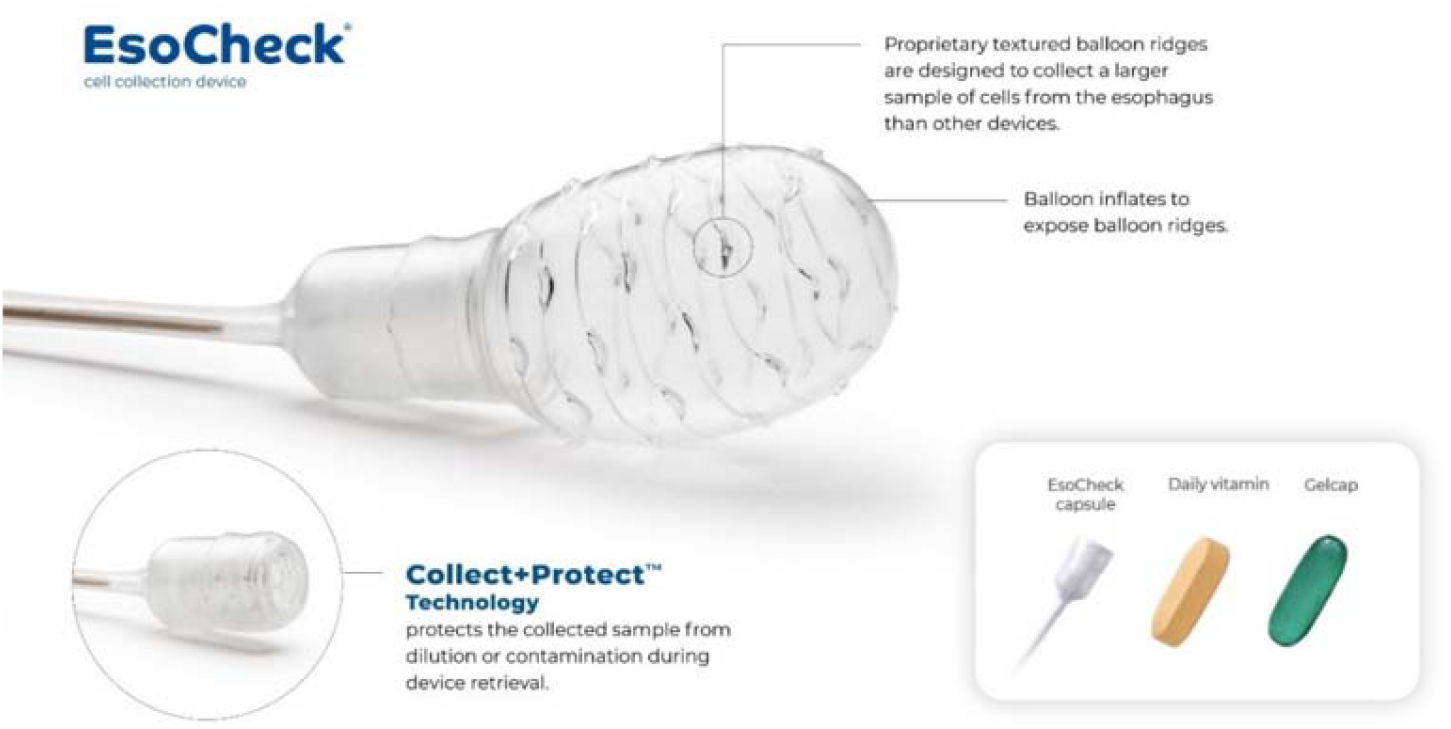
The EsoCheck® balloon capsule device including Collect + Protect™ technology.

### 2.3. EsoGuard Assay

EG is a laboratory-developed test that detects DNA methylation at 31 CpG sites on the Vimentin (*VIM*) and Cyclin A1 (*CCNA1*) genes. EG has been clinically validated to detect disease along the full BE progression spectrum, up to and including EAC, in four independent studies (two multi-center case-control and two prospective cohort studies) [14,15,17,18].

All EG testing was performed at LucidDx Labs (Lake Forest, CA), the clinical laboratory of Lucid Diagnostics, which is accredited by the College of American Pathologists (CAP), certified under the Clinical Laboratory Improvement Amendments (CLIA), and approved by the New York State Clinical Laboratory Evaluation Program (NY-CLEP). Following cell collection, EC samples underwent DNA extraction, bisulfite conversion, and polymerase chain reaction amplification in triplicate of targeted VIM and CCNA1 regions, followed by NGS and bioinformatic quantification of methylation across the 31 CpG sites. Each replicate is called positive if either methylated *VIM* (m*VIM*) or methylated *CCNA1* (m*CCNA1*) meets the validated cutoff, and the final EG result is assigned by majority rule (at least two of three replicates). This binary framework enhances interpretability and clinical actionability. The assay has been validated analytically in accordance with CAP, CLIA, and NY-CLEP requirements, as previously described [19].

A positive result indicates aberrant DNA methylation consistent with changes along the BE spectrum (metaplasia, dysplasia, or neoplasia) and should be followed by EGD for visual examination and staging, whereas a negative result indicates a low likelihood of BE or EAC. Samples with insufficient DNA or failed quality checks are reported as DNA Quantity Not Sufficient (QNS) or Unevaluable, respectively.

### 2.4. Outcome Measures

Outcome measures for EC include device tolerability, acceptability, and safety. Device tolerability was assessed using two metrics: (1) the proportion of successful cell collections compared to total attempted, and (2) the patient’s gag response score during sample collection. EC acceptability was assessed by optional, anonymous patient surveys issued after completion of the cell collection procedure. The surveys collected subjective measures of pre-procedure anxiety, ease of the cell collection, level of discomfort, whether the patient would recommend the test to others, and whether the patient would be willing to repeat the test if clinically indicated. Responses were recorded on a 10-point scale with 1 being the best and 10 the worst score.

EC safety was assessed through reported device malfunctions/failures and adverse events. Events were categorized according to class of malfunction or failure, and any consequences to patient health and welfare, if applicable. EG results were evaluated to assess potential population trends; the effect of individual risk factors as predictors for EG positivity were explored using predictive models.

### 2.5. Statistical Analysis

Summary statistics were performed for patient baseline characteristics and EC procedure characteristics. Means and standard deviations are presented where applicable. The Chi-square test was used for assessing differences between categorical variables.

Multivariable logistic regression models were constructed to develop a pred ctive algorithm for estimating the probability of a positive EG result. The model included the most relevant available BE/EAC risk factors: presence of symptoms associated with GERD, age >50, male sex, white race, smoking history, obesity (body mass index (BMI) ≥30), and family history of BE/EAC. To validate the model internally, resampling with bootstrap and optimism correction was performed [20], involving repeated sampling from the original dataset with replacement to create multiple datasets of the same size as the original. This approach allows for estimation of optimism in the training data by calculating the area under the curve 10,000 times using variations of the data through bootstrap resampling and comparing it with the original area under the curve. Odds ratios (OR) are reported to two decimal places and area under the curve (AUC) is reported to three decimal places; all other numbers are reported to a single decimal place.

## 3. Results

### 3.1. EC Performance and Acceptability

Between January 2023 and June 2024, a total of 11,991 patients underwent EC cell collection by Lucid Diagnostics personnel (Figure 2). Among these, 94.7% (11,355/11,991) successfully produced a cell sample for analysis. Cell collection failed due to gagging in 5.3% of patients (n=636). EC success rates were highly consistent over time, with quarterly successful cell collection rates ranging from 93.5% to 97.1%. Technical success rates did not vary significantly from month to month, or by individual sample collectors. Newly hired staff were noted to reach technical success rates matching their steady state almost immediately after completion of training.

**Figure 2.**
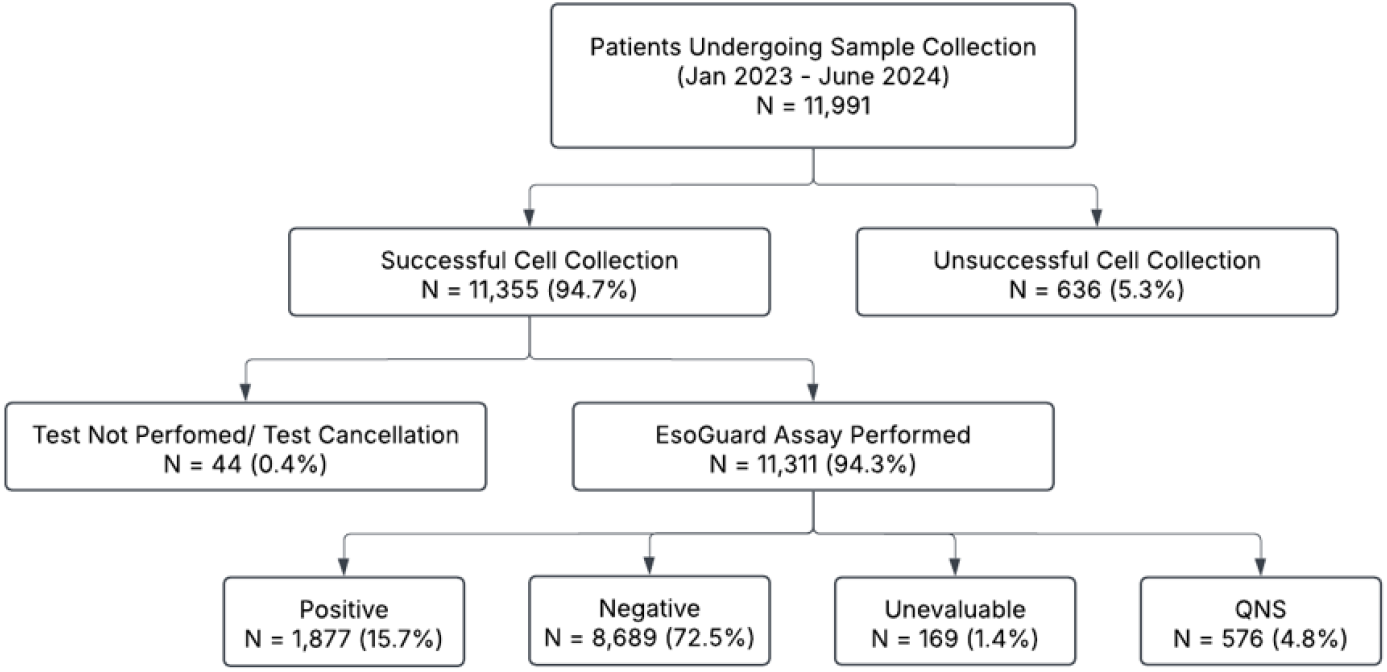
Flow diagram of patients undergoing EC cell sampling by Lucid personnel from January 2023 to June 2024.

Patient demographics and BE risk factors were analyzed for patients who successfully provided cell samples to the laboratory (N=11,355; Table 1). The most prevalent risk factors were white race (72.9%) and male sex (68.8%).

**Table 1.**
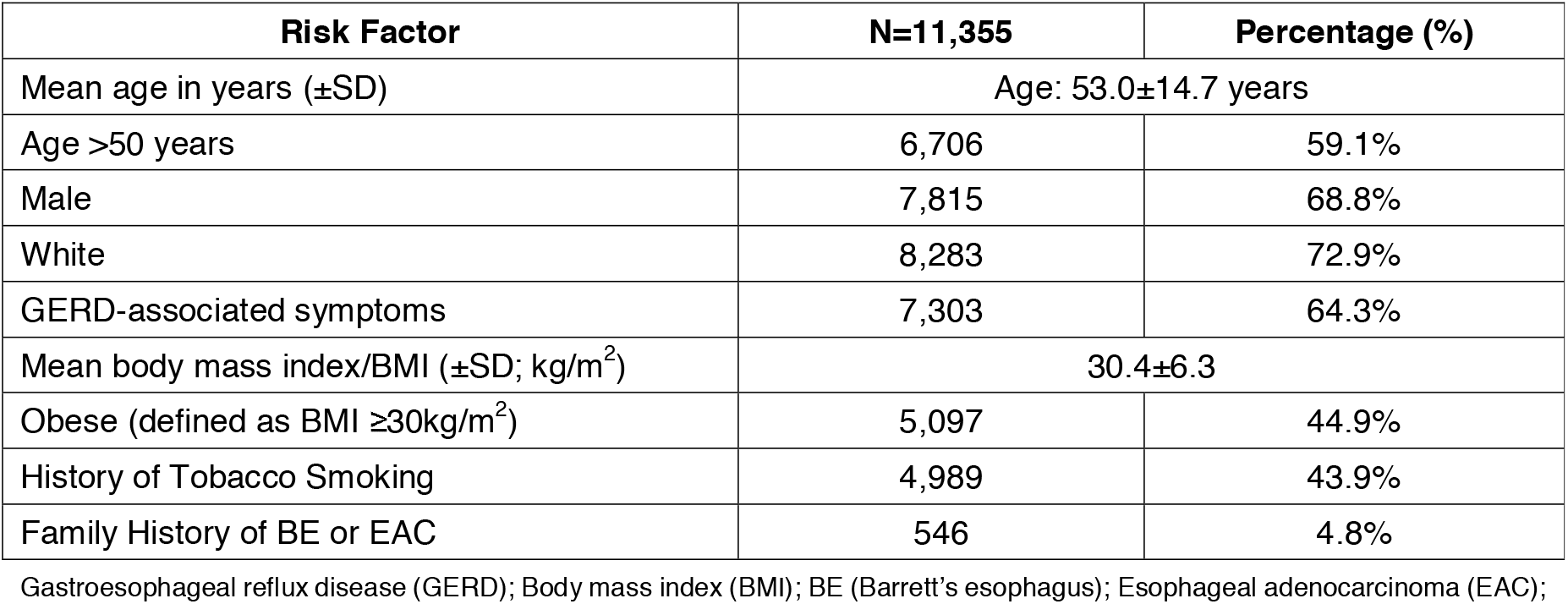
Baseline Characteristics for Patients with Successful EsoCheck Cell Collection.

Among patients who provided a cell sample, most (89.2%; 10,130/11,355) experienced no more than minimal to moderate gagging during cell collection (GR score ≤3). The average cell collection time was 1.9±0.9 minutes, and 98% (11,131/11,355) of cell collections were completed in less than 5 minutes. Satisfaction surveys were obtained from 586 patients (Appendix Supplemental Table 2). While most patients expressed at least some degree of pre-procedure anxiety, 82.4% of survey respondents reported low levels of discomfort (score ≤3) and 82.3% reported minimal difficulty swallowing the device (difficulty score ≤3). Additionally, 95.6% of respondents stated they would recommend EC to others in need of BE screening, and 94.6% would be willing to repeat the test if clinically indicated.

### 3.2. EC Device Complaints/Malfunctions and Safety

Device-related issues occurred in only 0.2% of patients, none of which resulted in patient harm (Table 2). The majority of device failures (22/29; 75.9%) were related to incomplete balloon encapsulation on removal, which was managed with repeat cell collection using a new device. No major device failures have been reported to date, including no instances of capsule detachment from the silicone tether.

**Table 2.** Device Complaints/Malfunctions Reported in the Quality Management System from Jan 2023 to Jun 2024.

| Failure Mode | Quantity | % (n/N)<br>N=11,991 |
| --- | --- | --- |
| Hole in Catheter* | 1 | 0.01% |
| Balloon Burst* | 1 | 0.01% |
| Balloon Microtear | 2 | 0.02% |
| Balloon did not fully encapsulate on removal | 22 | 0.18% |
| Syringe Sticking* | 1 | 0.01% |
| Missing Stopcock* | 1 | 0.01% |
| Deformed Stopcock* | 1 | 0.01% |

\*Device failures identified prior to patient exposure; in these instances, the cell collection was performed with a replacement device. As part of standard operating procedures, Lucid personnel always perform EsoCheck function checks prior to device administration. This allows identification of malfunctions and replacement of most devices before patient exposure

### 3.3. EG Results

Among 11,355 cell samples collected, binary test results were issued for 10,566 (93.1%; Figure 2). Reasons for laboratory non-results included: (a) 44 samples (<1%) that could not be analyzed either due to sample integrity issues, or test cancellation by the provider; (b) 576 samples (5.1%) that had insufficient DNA for analysis (reported as QNS); and (c) 169 samples (1.5%) that failed quality check (reported as Unevaluable). The EG positivity rate for all patients who attempted testing was 15.7% (1,877/11,991); the positivity rate among laboratory samples with first-pass yield was 17.8% (1,877/10,566).

### 3.4. Predictors of EG Positivity

A multivariable logistic regression analysis was performed to evaluate individual BE risk factors as predictors of EG positivity (Table 3). Age >50 years was shown to be the most predictive risk factor, with an OR of 2.37 (95% CI: 2.20–2.54, p-value <0.0001). EG positivity rates increased with age across the full analysis population (Figure 3). The presence of a family history of BE/EAC (OR: 1.19, 95% CI: 1.07–1.33, p-value = 0.0020) and tobacco smoking history (OR: 1.15, 95% CI: 1.10–1.21, p-value = <0.0001) also were associated with EG positivity, whereas obesity (BMI ≥30) was inversely associated with EG positivity (OR: 0.90, 95% CI: 0.86–0.95, p-value = 0.0001).

**Table 3.**
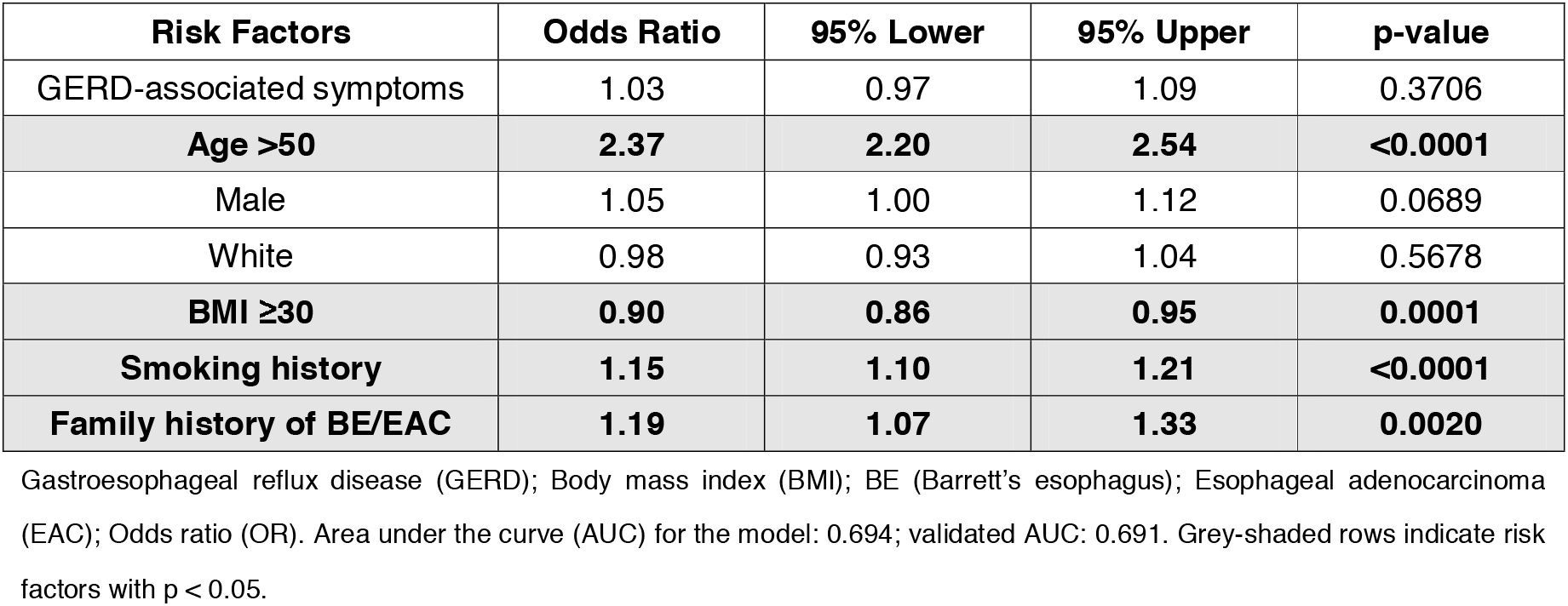
Risk Factor Prediction Model for EsoGuard Positivity Utilizing Multivariable Logistic Regression.

| Risk Factors | Odds Ratio | 95% Lower | 95% Upper | p-value |
| --- | --- | --- | --- | --- |
| GERD-associated symptoms | 1.03 | 0.97 | 1.09 | 0.3706 |
| **Age >50** | **2.37** | **2.20** | **2.54** | **<0.0001** |
| Male | 1.05 | 1.00 | 1.12 | 0.0689 |
| White | 0.98 | 0.93 | 1.04 | 0.5678 |
| **BMI ≥30** | **0.90** | **0.86** | **0.95** | **0.0001** |
| **Smoking history** | **1.15** | **1.10** | **1.21** | **<0.0001** |
| **Family history of BE/EAC** | **1.19** | **1.07** | **1.33** | **0.0020** |

Gastroesophageal reflux disease (GERD); Body mass index (BMI); BE (Barrett's esophagus); Esophageal adenocarcinoma (EAC); Odds ratio (OR). Area under the curve (AUC) for the model: 0.694; validated AUC: 0.691. Grey-shaded rows indicate risk factors with p < 0.05.

**Figure 3.**
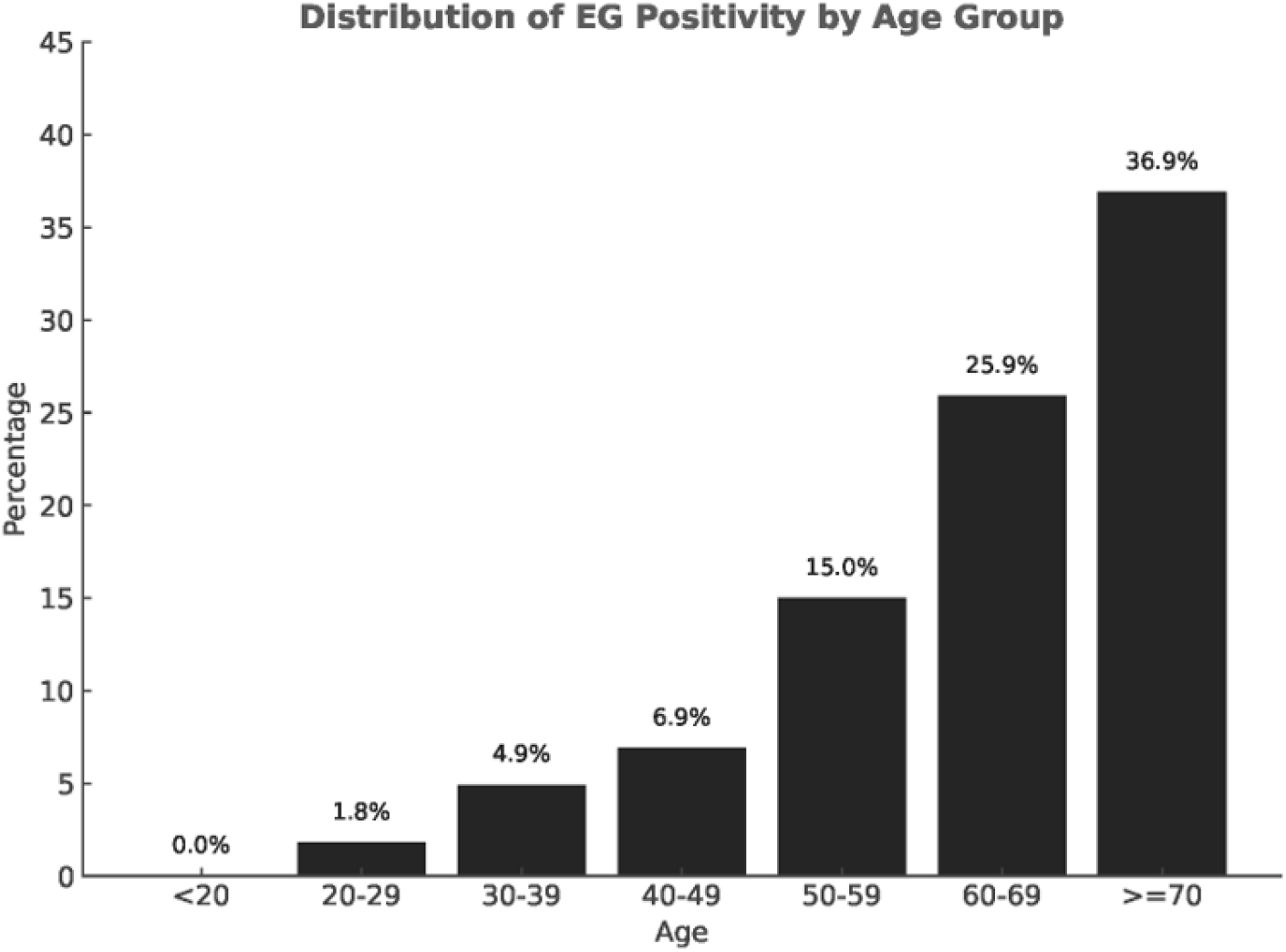
Rate of EsoGuard positivity across different patient age groups (in years) among the commercially tested population.

## 4. Discussion

This real-world data analysis represents the largest feasibility and tolerability study of non-endoscopic BE screening with EG/EC, encompassing nearly 12,000 patients. These data support rapid sample collection, safety, and tolerability of the EsoCheck device. Binary EG results were issued for 10,566 of the 11,355 successfully collected samples (93.1%), representing 88.1% of the 11,991 patients in whom collection was attempted. This finding suggests that EG/EC provides a practical and feasible non-endoscopic in-office approach to EGD triage for BE diagnosis.

EC’s balloon-capsule design with a durable silicone tether distinguishes it from other non-endoscopic cell collection devices such as Cytosponge™ (Medtronic, Galway, Ireland), EsophaCap® (Lucid Diagnostics Inc., New York, NY), and EndoSign® (Cyted Ltd, Cambridge, UK), which use sponge-on-a-string (SoS) based technology. These SoS devices consist of mesh spheres compressed within gelatin capsules that expand upon deployment and are withdrawn through the proximal digestive tract. This approach results in non-targeted, pan-esophageal sampling, collecting surface mucosal cells as the sponge is withdrawn from the stomach, the entire length of the esophagus, and the oropharynx. SoS devices typically require approximately 10 min for sample collection, as the gelatin capsule must dissolve in the stomach for at least seven minutes before retrieval [21–23]. In addition, reports of sponge detachment during retrieval, necessitating endoscopic removal, have led to FDA recalls of both the Cytosponge™ and EsophaCap® devices [21,22,24,25]. In contrast, the present real-world dataset demonstrates that EC sample collection is completed in under five minutes in 98% of cases, with an overall administration success rate of 95%, comparable or even superior to other non-endoscopic collection devices [22–24,26]. This rapid administration facilitates use in high-throughput settings, such as health fairs, and allows for easier integration into busy outpatient practice workflows. Importantly, with no reported device-related adverse events and only a 0.2% incidence of any device malfunctions, EC demonstrates a highly favorable safety profile.

The analytical and clinical performance characteristics of the EG/EC test (including sensitivity, specificity, positive predictive value [PPV], and negative predictive value [NPV]) have been rigorously established and previously were published in independent clinical validation studies. This study is intended to complement those prior efforts by demonstrating feasibility, adoption, and performance of the testing paradigm in real-world clinical workflows. It is notable that in this real-world experience, 17.8% of laboratory samples with first-pass yield were EG positive, aligning with a previously published prospective single-arm screening study that reported a 19% EG positivity rate and 8% BE prevalence on confirmatory EGD [15].

Age >50 was a strong predictor of EG positivity (OR: 2.37) on multivariable logistic regression models. This aligns with published literature demonstrating higher BE prevalence in older age groups (most notably among those aged >60 years) [27]. Tobacco smoking and family history of BE or EAC also were predictors of EG positivity, with respective odds ratios of 1.15 and 1.19. However, these values were lower than expected, as prior studies report odds ratios up to 12.2 for family history and 1.8 for SIM among smokers [28,29]. This discrepancy may stem from inaccurate risk factor reporting, as patients often lack familiarity with full family medical history, and literature suggests about 8% withhold smoking status from their healthcare providers, potentially due to perceived stigma [30]. Obesity (BMI ≥30) was also unexpectedly inversely associated with EG positivity (OR: 0.90; 95% CI: 0.86–0.95), despite being an established risk factor for BE and EAC. However, among patients with GERD, BMI has shown little independent predictive value for BE [31], and central adiposity, rather than overall BMI, appears to drive this risk [32]. BMIs calculated from inaccurate or outdated patient-reported height and weight may also have contributed to this finding.

The finding that the presence of GERD symptoms was not a statistically significant predictor of EG positivity was unexpected, given that reflux is the presumed underlying mechanism driving BE metaplasia. However, a growing body of literature suggests that symptoms may be a poor indicator for the presence of chronic GERD. Studies have shown that a substantial proportion of asymptomatic individuals may have esophagitis, and up to 50% of patients who ultimately develop esophageal adenocarcinoma do not report frequent GERD symptoms beforehand [33,34]. Reliance on patient self-reporting of GERD symptoms also may have contributed to the lack of apparent association with EG positivity. While heartburn commonly is identified as a manifestation of GERD, atypical or extra-esophageal symptoms may be underrecognized or dismissed. Conversely, intermittent or isolated episodes of heartburn may be overreported as GERD in the absence of chronic or clinically meaningful disease. Either scenario could have attenuated any true association between GERD symptoms and EG positivity.

Key study limitations include both the retrospective, observational study design and the reliance on self-reported risk factors, which may introduce misclassification and social desirability bias (e.g., GERD, smoking, BMI) despite the large sample size. Limiting the cohort to patients tested by Lucid Diagnostics personnel also may have introduced selection bias. However, as more than 80% of total commercial tests performed during the study period were conducted by the same employed personnel, the findings remain broadly representative of the commercial testing population. Additionally, confirmatory EGD and histopathology results are not captured in the laboratory database, and therefore not directly available for the patients in this study. EGD referral is managed by the patient’s ordering provider and outside the scope of laboratory services. As such, the proportion of EG-positive patients who adhered to recommended EGD, and those with confirmed BE or EAC on histopathology could not be determined in this real-world cohort. To address this gap, an ongoing registry (ClinicalTrials.gov identifier: NCT05965999) is prospectively tracking EGD and histopathology outcomes, and these findings will be reported in a separate manuscript. The diagnostic performance of the test should be interpreted in the context of prior clinical validation studies [14,15,17,18]. Finally, patient acceptability data were derived from a voluntary, anonymous survey completed by only 586 patients (5.2% of those with successful cell collection). These responses may be subject to self-selection and non-response bias and may not fully represent the experience of the broader tested population.

## 5. Conclusions

In conclusion, this study represents the largest real-world evaluation of non-endoscopic BE testing using EG/EC to date. The EC device was well tolerated, safe, and efficient, supporting its use in high-volume settings. EG results generally were consistent with the population risk profile. Logistic regression identified age >50 as a strong predictor of test positivity, likely reflecting increased disease prevalence. Overall, these findings support the feasibility of EG/EC for large-scale implementation and highlight their potential to expand access to BE screening in high-risk patients across diverse healthcare settings.

## Data Availability

All data produced in the present study are available upon reasonable request to the authors.

## Author Contributions

Conceptualization, I.T.L., S.V. and V.T.L.; methodology, I.T.L., S.V. and V.T.L.; formal analysis, I.T.L.; investigation, I.T.L., S.V. and V.T.L.; data curation, I.T.L. and V.T.L.; writing—original draft preparation, I.T.L., S.V. and V.T.L.; writing—review and editing, M.S.S., W.T.C., P.S.P. and L.A.; visualization, I.T.L. All authors have read and agreed to the published version of the manuscript.

## Institutional Review Board Statement

This study protocol was approved by the Western Institutional Review Board (WIRB®; study number 20244309), with the Board finding the study meets requirements for waivers of consent under 21 CFR 50.22.

## Informed Consent Statement

Patient consent was waived by the Western Institutional Review Board under 21 CFR 50.22.

## Data Availability Statement

Aggregate data supporting the findings of this study may be made available from the corresponding author upon reasonable request, subject to a data use agreement and applicable privacy regulations.

## Abbreviations

The following abbreviations are used in this manuscript:

BE: Barrett’s Esophagus
SIM: Specialized Intestinal Metaplasia
EAC: Esophageal Adenocarcinoma
EG: EsoGuard
EC: EsoCheck
GERD: Gastroesophageal Reflux Disease
EGD: Esophagogastroduodenoscopy
ACG: American College of Gastroenterology
AGA: American Gastroenterological Association
NGS: Next-Generation Sequencing
*VIM*: Vimentin
*CCNA1*: Cyclin A1
m*VIM*: Methylated Vimentin
m*CCNA1*: Methylated Cyclin A1
PPV: Positive Predictive Value
NPV: Negative Predictive Value
QNS: Quantity Not Sufficient

## Appendix

**Appendix Supplemental Table 1.** Gag Response Scoring.

| Score | Severity of gag response | Detailed description |
| --- | --- | --- |
| 1 | None to minimal | No gagging or patient only gagged during the initial swallow |
| 2 | Mild | Non-forceful, infrequent gagging during the procedure (occurring a few times but not through-out the entirety) |
| 3 | Moderate | Non-forceful gagging through-out the procedure |
| 4 | Severe but able to complete cell collection | Frequent gagging with intense forceful retch, but still able to complete the cell collection |
| 5 | Unable to complete cell collection | Unable to complete the cell collection whether due to retching or not |

**Appendix Supplemental Table 2.**
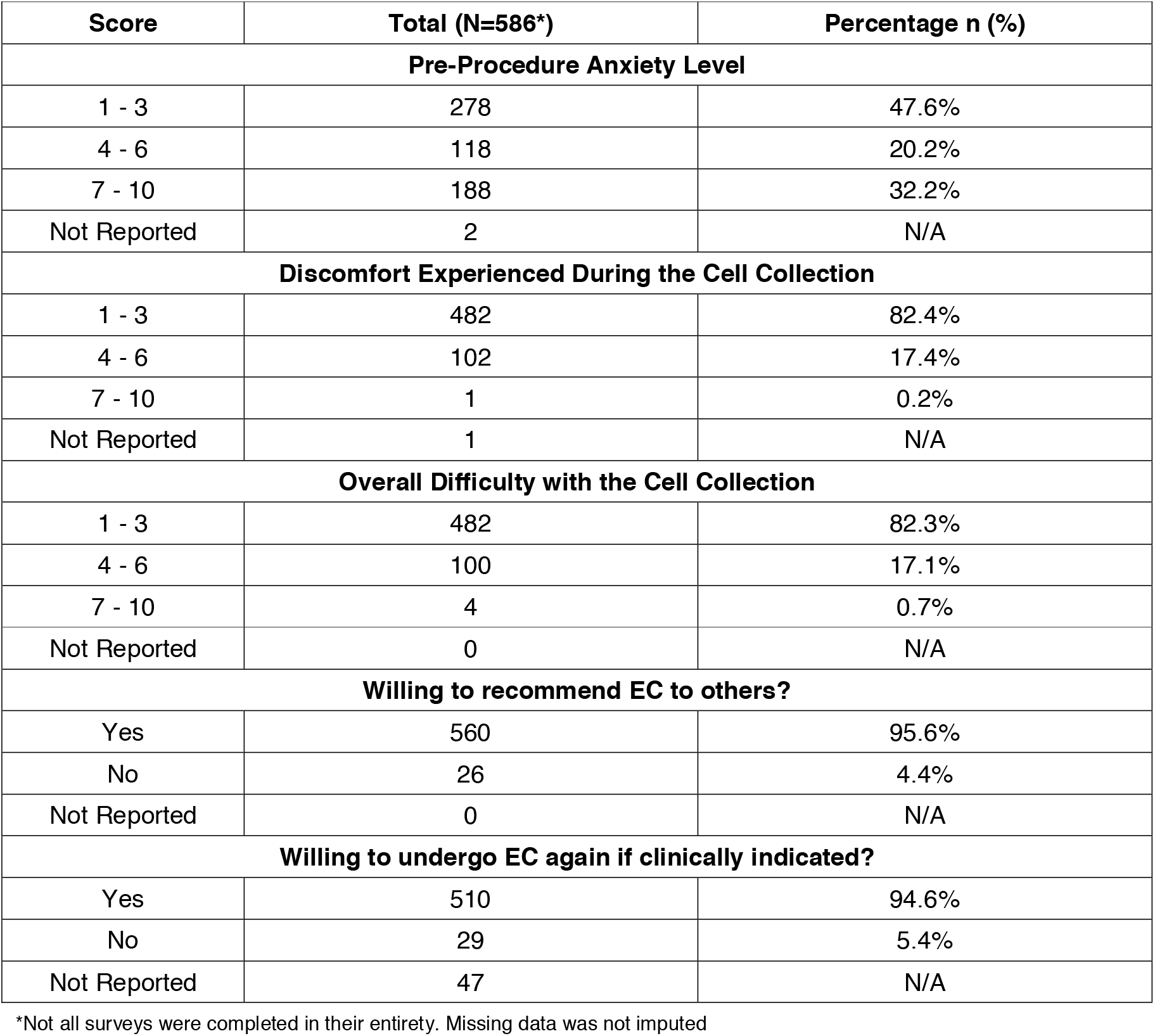
Results of the Patients Satisfactory Survey.

| Score | Total (N=586*) | Percentage n (%) |
| --- | --- | --- |
| **Pre-Procedure Anxiety Level** |  |  |
| 1 - 3 | 278 | 47.6% |
| 4 - 6 | 118 | 20.2% |
| 7 - 10 | 188 | 32.2% |
| Not Reported | 2 | N/A |
| **Discomfort Experienced During the Cell Collection** |  |  |
| 1 - 3 | 482 | 82.4% |
| 4 - 6 | 102 | 17.4% |
| 7 - 10 | 1 | 0.2% |
| Not Reported | 1 | N/A |
| **Overall Difficulty with the Cell Collection** |  |  |
| 1 - 3 | 482 | 82.3% |
| 4 - 6 | 100 | 17.1% |
| 7 - 10 | 4 | 0.7% |
| Not Reported | 0 | N/A |
| **Willing to recommend EC to others?** |  |  |
| Yes | 560 | 95.6% |
| No | 26 | 4.4% |
| Not Reported | 0 | N/A |
| **Willing to undergo EC again if clinically indicated?** |  |  |
| Yes | 510 | 94.6% |
| No | 29 | 5.4% |
| Not Reported | 47 | N/A |

\*Not all surveys were completed in their entirety. Missing data was not imputed

## Disclaimer/Publisher’s Note

The statements, opinions and data contained in all publications are solely those of the individual author(s) and contributor(s) and not of MDPI and/or the editor(s). MDPI and/or the editor(s) disclaim responsibility for any injury to people or property resulting from any ideas, methods, instructions or products referred to in the content.

## References

1. Codipilly DC, Sawas T, Dhaliwal L, Johnson ML, Lansing R, Wang KK, et al. Epidemiology and Outcomes of Young-Onset Esophageal Adenocarcinoma: An Analysis from a Population-Based Database. Cancer Epidemiol Biomarkers Prev. 2021;30(1):142–9.

2. National Cancer Institute: Surveillance E, and End Results Program. Esophageal Cancer — Cancer Stat Facts 2026 [Available from: https://seer.cancer.gov/statfacts/html/esoph.html.]

3. Curtius K, Rubenstein JH, Chak A, Inadomi JM. Computational modelling suggests that Barrett’s oesophagus may be the precursor of all oesophageal adenocarcinomas. Gut. 2020;70(8):1435–40.

4. Zamani SA, Wu L, Black EL, Bartram A, Ng AWT, Secrier M, et al. Integrated epidemiological and molecular data inform the relationship between precancer and cancer states of esophageal adenocarcinoma. Nat Med. 2026;32(5):1805–16.

5. Shaheen NJ, Falk GW, Iyer PG, Souza RF, Yadlapati RH, Sauer BG, et al. Diagnosis and Management of Barrett’s Esophagus: An Updated ACG Guideline. Am J Gastroenterol. 2022;117(4):559–87.

6. Muthusamy VR, Wani S, Gyawali CP, Komanduri S. AGA Clinical Practice Update on New Technology and Innovation for Surveillance and Screening in Barrett’s Esophagus: Expert Review. Clin Gastroenterol Hepatol. 2022;20(12):2696-706.e1.

7. Kamboj AK, Katzka DA, Iyer PG. Endoscopic Screening for Barrett’s Esophagus and Esophageal Adenocarcinoma: Rationale, Candidates, and Challenges. Gastrointest Endosc Clin N Am. 2021;31(1):27–41.

8. Eluri S, Reddy S, Ketchem CC, Tappata M, Nettles HG, Watts AE, et al. Low Prevalence of Endoscopic Screening for Barrett’s Esophagus in a Screening-Eligible Primary Care Population. Am J Gastroenterol. 2022;117(11):1764–71.

9. Kolb JM, Chen M, Tavakkoli A, Gallegos J, O’Hara J, Tarter W, et al. Patient Knowledge, Risk Perception, and Barriers to Barrett’s Esophagus Screening. Am J Gastroenterol. 2023;118(4):615–26.

10. Peery AF, Dellon ES, Lund J, Crockett SD, McGowan CE, Bulsiewicz WJ, et al. Burden of gastrointestinal disease in the United States: 2012 update. Gastroenterology. 2012;143(5):1179-87.e3.

11. Menezes A, Tierney A, Yang YX, Forde KA, Bewtra M, Metz D, et al. Adherence to the 2011 American Gastroenterological Association medical position statement for the diagnosis and management of Barrett’s esophagus. Dis Esophagus. 2015;28(6):538–46.

12. Runge TM, Abrams JA, Shaheen NJ. Epidemiology of Barrett’s Esophagus and Esophageal Adenocarcinoma. Gastroenterol Clin North Am. 2015;44(2):203–31.

13. Administration USFaD. 510(k) Premarket Notification K230339 2023 [Available from: https://www.accessdata.fda.gov/scripts/cdrh/cfdocs/cfpmn/pmn.cfm?ID=k230339

14. Moinova HR, LaFramboise T, Lutterbaugh JD, Chandar AK, Dumot J, Faulx A, et al. Identifying DNA methylation biomarkers for non-endoscopic detection of Barrett’s esophagus. Sci Transl Med. 2018;10(424).

15. Shaheen NJ, Othman MO, Taunk J, Chang KJ, Jaganmohan S, Yachimski PS, et al. Use of a Two-Gene Methylated DNA Biomarker Assay and Nonendoscopic Balloon for Detection of Barrett Esophagus Among High-Risk Individuals in a Screening Population. Am J Gastroenterol. 2024;120(7):1511–20.

16. Lister D, Fine A, Maheshwari S, Bradley PS, Lister K, Lee VT, et al. Real-World Clinical Utility of a Methylated DNA Biomarker Assay on Samples Collected with a Swallowable Capsule-Balloon for Detection of Barrett’s Esophagus (BE). Medicina. 2024;60(12):2052.

17. Moinova HR, Verma S, Dumot J, Faulx A, Iyer PG, Canto MI, et al. Multicenter, Prospective Trial of Nonendoscopic Biomarker-Driven Detection of Barrett’s Esophagus and Esophageal Adenocarcinoma. Am J Gastroenterol. 2024;119(11):2206–14.

18. Greer KB, Blum AE, Faulx AL, Deming EM, Hricik LL, Siddiqui H, et al. Nonendoscopic Screening for Barrett’s Esophagus and Esophageal Adenocarcinoma in At-Risk Veterans. Am J Gastroenterol. 2025;120(3):545–53.

19. Ghosal A, Verma S, Le IT, Lee VT, deGuzman BJ, Aklog L. Analytical Validation of a DNA Methylation Biomarker Test for the Diagnosis of Barrett’s Esophagus and Esophageal Adenocarcinoma from Samples Collected Using EsoCheck(®), a Non-Endoscopic Esophageal Cell Collection Device. Diagnostics (Basel). 2024;14(16).

20. Efron B, & Tibshirani, R.J. An Introduction to the Bootstrap. 1 ed. New York: Chapman and Hall/CRC; 1994.

21. Shaheen NJ, Komanduri S, Muthusamy VR, Wani S, O’Donovan M, Kaushal R, et al. Acceptability and Adequacy of a Non-endoscopic Cell Collection Device for Diagnosis of Barrett’s Esophagus: Lessons Learned. Dig Dis Sci. 2022;67(1):177–86.

22. Iyer PG, Slettedahl SW, Mahoney DW, Giakoumopoulos M, Olson MC, Krockenberger M, et al. Algorithm Training and Testing for a Non-Endoscopic Barrett’s Esophagus Detection Test in Prospective Multicenter Cohorts. Clin Gastroenterol Hepatol. 2024.

23. Ross-Innes CS, Debiram-Beecham I, O’Donovan M, Walker E, Varghese S, Lao-Sirieix P, et al. Evaluation of a minimally invasive cell sampling device coupled with assessment of trefoil factor 3 expression for diagnosing Barrett’s esophagus: a multi-center case-control study. PLoS Med. 2015;12(1):e1001780.

24. Fitzgerald RC, di Pietro M, O’Donovan M, Maroni R, Muldrew B, Debiram-Beecham I, et al. Cytosponge-trefoil factor 3 versus usual care to identify Barrett’s oesophagus in a primary care setting: a multicentre, pragmatic, randomised controlled trial. Lancet. 2020;396(10247):333–44.

25. Januszewicz W, Tan WK, Lehovsky K, Debiram-Beecham I, Nuckcheddy T, Moist S, et al. Safety and Acceptability of Esophageal Cytosponge Cell Collection Device in a Pooled Analysis of Data From Individual Patients. Clin Gastroenterol Hepatol. 2019;17(4):647-56.e1.

26. Iyer PG, Taylor WR, Johnson ML, Lansing RL, Maixner KA, Hemminger LL, et al. Accurate Nonendoscopic Detection of Barrett’s Esophagus by Methylated DNA Markers: A Multisite Case Control Study. Am J Gastroenterol. 2020;115(8):1201–9.

27. Vissapragada R, Bulamu NB, Whiteman DC, Bright T, Karnon J, Watson DI. Computing lifetime incidence of esophageal adenocarcinoma and age-specific prevalence of Barrett’s esophagus. Dis Esophagus. 2025;38(3).

28. Chak A, Lee T, Kinnard MF, Brock W, Faulx A, Willis J, et al. Familial aggregation of Barrett’s oesophagus, oesophageal adenocarcinoma, and oesophagogastric junctional adenocarcinoma in Caucasian adults. Gut. 2002;51(3):323–8.

29. Edelstein ZR, Bronner MP, Rosen SN, Vaughan TL. Risk factors for Barrett’s esophagus among patients with gastroesophageal reflux disease: a community clinic-based case-control study. Am J Gastroenterol. 2009;104(4):834–42.

30. Stuber J, Galea S. Who conceals their smoking status from their health care provider? Nicotine Tob Res. 2009;11(3):303–7.

31. Cook MB, Greenwood DC, Hardie LJ, Wild CP, Forman D. A systematic review and meta-analysis of the risk of increasing adiposity on Barrett’s esophagus. Am J Gastroenterol. 2008;103(2):292–300.

32. Singh S, Sharma AN, Murad MH, Buttar NS, El-Serag HB, Katzka DA, et al. Central adiposity is associated with increased risk of esophageal inflammation, metaplasia, and adenocarcinoma: a systematic review and meta-analysis. Clin Gastroenterol Hepatol. 2013;11(11):1399-412.e7.

33. Du Y, Spencer A, Roy P, Abdulhamid A, Dalessio S, Levenick J, et al. Asymptomatic Erosive Esophagitis. Dig Dis Sci. 2025;70(2):462–8.

34. Nguyen TH, Thrift AP, Rugge M, El-Serag HB. Prevalence of Barrett’s esophagus and performance of societal screening guidelines in an unreferred primary care population of U.S. veterans. Gastrointest Endosc. 2021;93(2):409-19.e1.

